# Clinical outcomes and mortality risk among inborn and referred newborns admitted to hospitals in Kenya

**DOI:** 10.64898/2026.03.03.26347492

**Authors:** Judy Baariu, Sarah Murless-Collins, George Okello, Dolphine Mochache, Franklin Okech, Lucas Malla, James H. Cross, David Gathara, Joy E. Lawn, Eric O. Ohuma, William M. Macharia, Rebecca E. Penzias

## Abstract

**Background:** Newborns requiring inpatient care, particularly small and sick newborns (SSNBs), face high risk of mortality. Newborns referred from other facilities may experience worse outcomes than those born and managed within the same hospital (*inborn newborns*). Understanding factors contributing to this disparity in outcomes could support efforts to scale-up care and accelerate progress towards achieving Sustainable Development Goals target 3.2.

**Methods:** Data on 130,773 newborns admitted to 13 hospitals implementing with NEST360 in Kenya were obtained from the Neonatal Inpatient Dataset, between January 2019–October 2024. We described characteristics and primary diagnoses. Logistic regression was used to evaluate factors associated with mortality.

**Results:** Among admissions, 114,084 (87.2%) were inborn and 16,689 (12.8%) referred. Referred newborns were more likely to be extremely preterm (6.1% vs 3.1%), have extremely low birthweight (<1,000g) (4.6% vs 2.6%) and present with respiratory distress (26.2% vs 15.0%) and hypoxia (23.2% vs 15.3%) compared to those inborn. Only 59.6% of referred newborns were admitted on first day of life compared to 80.2% inborn newborns. Unadjusted mortality among referred newborns was 29.0% compared to 11.3% in those inborn. Risk factors associated with mortality among referred newborns included being extremely low birthweight (odds ratio [OR] 13.57, 95% CI 11.19–16.44), respiratory distress (OR 4.07, 95% CI 3.77–4.39), and congenital anomalies (OR 1.66, 95% CI 1.41–1.95). Prematurity and intrapartum-related complications were also associated with increased odds of death. In multivariable analysis, being referred remained strongly associated with mortality (adjusted OR [aOR] 2.54, 95% CI 2.39–2.71).

**Conclusion:** Referred newborns had nearly three times higher odds of mortality compared to those inborn. This may highlight referral selection bias amongst this group and could also be related to inadequate pre-referral stabilisation, unsafe neonatal transportation and admission delays. If successfully implemented, a strong hub-and-spoke approach may improve care at lower levels of care and decongest receiving facilities. Overall, improving quality of care across the continuum of referral process is a cornerstone in strategies to reduce neonatal mortality towards attainment of national and global newborn survival targets.

**KEY FINDINGS:** *1. WHAT WAS KNOWN?:*

- Neonatal mortality remains high in sub-Saharan Africa and newborns referred from other health facilities may experience poorer outcomes than those born and managed within the same hospital.
- There is limited evidence on morbidity and mortality outcomes among inborn and referred newborns. This is important to inform specialised newborn care and targeted improvements in referral.

*2. WHAT WAS DONE THAT IS NEW?:*

- This study analysed routinely collected clinical data on 130,773 newborns admitted to 13 hospitals implementing with NEST360 in Kenya between 2019 and 2024.
- Diagnoses outcomes and neonatal characteristics were described and compared between inborn and referred newborns. Factors associated with neonatal mortality were also examined using logistic regression analysis.

*3. WHAT WAS FOUND?:*

- Referred newborns had higher unadjusted mortality rate than inborn newborns (29.0% vs 11.3%; p<0.001), with 3 times higher odds of death in univariable logistic regression analysis (OR 3.20, 95% CI 3.08–3.33).
- Referred newborns were more clinically vulnerable at admission and had higher proportions of extreme prematurity (6.1% vs 3.1%), very preterm birth (14.0% vs 8.6%), and extremely low birthweight (4.6% vs 2.6%). Among both groups, key risk factors associated with mortality included birthweight, gestational age, respiratory distress, hypothermia, and clinical diagnoses.
- Among referred newborns some of the risk factors associated with mortality included being extremely low birthweight (OR 13.57, 95% CI 11.19–16.44), respiratory distress (OR 4.07, 95% CI 3.77–4.39), congenital anomalies (OR 1.66, 95% CI 1.41–1.95), and intrapartum-related complications (OR 1.35, 95% CI 1.20–1.52).

*4. WHAT NEXT?:*

- Strengthen neonatal referral systems through clearer referral criteria, improved pre-referral stabilisation, better neonatal transport, and prompt triage on arrival at receiving hospitals. Routine clinical data should be used to monitor referral processes and outcomes and to guide continuous quality improvement.
- Further research is needed to capture referral to admission time, transport characteristics, and quality of pre-referral stabilisation. Linking neonatal admission data with maternal records and assessing outcomes beyond hospital discharge would also improve understanding of referral pathways and long-term outcomes.

## BACKGROUND

The first 28 days of life remain the most vulnerable in the human lifespan, with nearly half of all under-five deaths occurring during this period. Progress towards achieving Sustainable Development Goal (SDG) target 3.2 – to end preventable newborn deaths by 2030 – has been slow in sub-Saharan Africa, which has the highest neonatal mortality rate of 26 deaths per 1,000 live births, more than twice the SDG aim of 12 per 1,000 [1, 2]. Newborns who require inpatient care including those born prematurely, with low birthweight, and/or who become sick (referred to as *small and sick newborns (SSNBs)*) are at increased risk of mortality and long-term complications [3–5]. Newborns referred from other facilities are particularly vulnerable to these poor outcomes compared with those born and managed within the same facility (*inborn newborns*). Referred SSNBs are prone to severe clinical conditions including respiratory distress, neonatal sepsis, birth asphyxia, and hypothermia [6–9]. Studies in sub-Saharan Africa have also shown that newborns born outside the admitting hospital have nearly twice the mortality compared to inborn newborns [8, 9]. In Kenya, a study reported an unadjusted mortality rate of 26.0% among newborns born outside three referral hospitals [10], compared with 10.2% among those inborn in a separate multisite study [11].

Many of these adverse outcomes are preventable but require very rapid action, sometimes within minutes. Neonatal mortality is therefore a sensitive marker of health systems performance and connections between levels of care [12]. Disparities in outcomes show the need to strengthen care throughout the referral pathway and to better understand factors contributing to worse outcomes among referred newborns. The World Health Organization provides standards for improving quality of care to help vulnerable babies survive and thrive, with functioning referral systems identified as a key component to ensure timely and appropriate transfer to higher-level facilities for specialised care when needed [13]. In addition, the Every Woman Every Newborn Everywhere (EWENE) initiative [14] and Emergency Obstetric and Newborn Care (EmONC) referral signal functions [15] include recommendations to strengthen health worker capacities, transport systems, referral mechanisms, and networks of health facilities.

While studies in low- and middle-income countries (LMICs) have shown that timely identification of newborns requiring referral and appropriate facilitation of the process can reduce mortality and morbidity [16–18], there are persistent health care system challenges. These include inadequate pre-referral stabilisation, lack of clear referral protocols, limited communication between facilities, and delays in decision-making at lower levels of care [19, 20]. Transportation barriers – such as long distances [21, 22], limited neonatal transport readiness, lack of essential medical supplies [10], and shortage of trained personnel during transfer [6] – may further lead to adverse outcomes by disrupting continuity of care and delaying life-saving interventions. At receiving facilities, overcrowding and shortage of staff or essential supplies may also limit capacity to provide immediate and specialised care [23, 24]. Assessing newborn outcomes therefore provides an opportunity to identify factors that may be associated with gaps in care and inform need for targeted interventions for example during referral and admission processes [25].

### Aim and objectives

In this paper, we aim to compare clinical outcomes and mortality risk among inborn and referred newborns admitted to hospitals implementing with NEST360 in Kenya. The specific objectives were to:

1. Describe characteristics of inborn and referred newborns.
2. Describe clinical diagnoses during hospitalisation among inborn and referred newborns.
3. Evaluate factors associated with mortality among inborn and referred newborns.

## METHODS

### Study setting

NEST360 was established in 2019 to support African governments to scale-up small and sick newborn care (SSNC) by implementing a health systems strengthening package, including evidence-based care using locally-owned data [26]. During Phase 1, the programme was implemented in 66 hospitals across Kenya, Malawi, Nigeria, and Tanzania through to the end of 2023. In 2024, NEST360 expanded to Phase 2, including Ethiopia and additional hospitals in Kenya, Nigeria, and Tanzania. This analysis used data from 13 Phase 1 hospitals implementing with NEST360 across 10 of the 47 counties in Kenya.

### Data source

Individual-level data were collected using a de-identified Neonatal Inpatient Dataset (NID) integrated within the Clinical Information Network, which is an established neonatal and paediatric data collection platform in Kenya [27, 28]. Information on each newborn admitted to the implementing hospitals was routinely gathered from patient forms and clinical case notes and entered by trained on-site data collectors using REDCap [29].

Data collected between January 2019–October 2024 were analysed. The dataset included key variables across five domains: birth details and maternal history, admission and referral information (including birth location, reason for referral and admission), clinical observations, nursing care, and discharge outcomes. All inborn newborns and those referred from other facilities and admitted to the 13 hospitals during the study period were eligible for inclusion. Newborns born at home were excluded from analysis.

### Methods by objectives

#### Objective 1: Describe characteristics of inborn and referred newborns

Newborn admission records were reviewed to determine birth location (place of birth) and to assess completeness of data on age. Records with missing data on birth location or age, those born outside a health facility, and babies older than 28 days at admission were excluded from the analysis. Inborn newborns and those referred from other health facilities were retained for the final analysis.

Newborn characteristics were categorised as demographic, perinatal, clinical, and maternal. Frequencies and percentages were reported for categorical variables while median and interquartile ranges (IQRs) were reported for continuous variables.

#### Objective 2: Describe clinical diagnoses among inborn and referred newborns

The newborns primary diagnosis and other clinical diagnoses were recorded. These were grouped into nine categories for analysis: congenital anomalies, prematurity, infection, intra-partum related complications, pathological jaundice, macrosomia, tachypnoea, term newborns with low birthweight (LBW) (<1,500g), and other diagnoses. Diagnoses categories were then disaggregated by birth location (inborn and referred). The number and proportion of diagnoses across groups was calculated and visualised using a catplot.

#### Objective 3: Compare factors associated with neonatal mortality among inborn and referred newborns

To evaluate factors associated with neonatal mortality, univariable logistic regression was first conducted for each potential predictor, stratified by place of birth (inborn and referred). Variables assessed included place of birth, sex, birthweight, gestational age, small vulnerable newborn category, respiratory distress, admission temperature, mode of delivery, singleton birth, diagnoses, maternal age, maternal survival status, and maternal HIV status. To define small vulnerable newborn categories, newborns were classified into four groups based on gestational age (preterm <37 weeks vs term ≥37 weeks), and size for gestational age (small for gestational age (SGA) <10th centile vs non-SGA ≥10th centile). These included term non-SGA (reference), term SGA, preterm non-SGA, and preterm SGA. Size for gestational age was defined using INTERGROWTH-21^st^ newborn standards, with SGA defined as birthweight below 10th centile for gestational age and sex [30, 31]. Diagnosis categories with very low frequency and/or minimal mortality were excluded from regression analyses to ensure meaningful interpretation. These included tachypnoea, macrosomia, other LBW diagnoses among term newborns, and the heterogeneous ‘other’ diagnosis category.

Variables with a p-value <0.25 in univariable analysis were selected for inclusion in multivariable logistic regression. The analysis was performed using forward stepwise selection, with a p-value cut-off point of 0.25 for variable entry (pe=0.25). This slightly higher cut-off was selected to avoid excluding variables that may have important adjusted associations or act as confounders in the multivariable model [32]. Variables with very small numbers in some categories or limited variation in outcome were automatically omitted by the model to ensure reliable estimation. Adjusted odds ratios (aORs) with 95% confidence intervals (CIs) were reported with p-values. All data cleaning, analyses, and visualisations were conducted using Stata version 19.

## RESULTS

A total of 147,770 newborn records were reviewed, and 1,724 (1.2%) newborns born at home and 13,574 (9.2%) with missing or unspecified data on birth location excluded. An additional 1,699 (1.3%) with missing data on age and those older than 28 days were removed. Data on 130,773 newborns were retained, including 114,084 (87.2%) inborn and 16,689 (12.8%) referred newborns. **Additional File 1** summarises the sample inclusion process and distribution across 13 hospitals.

### Results by objectives

#### Objective 1: Newborn characteristics

**Table 1** presents baseline characteristics of 130,773 newborns. Most admissions occurred within the first day of life (77.6%), more commonly among inborn (80.2%) than referred newborns (59.6%, p<0.001). Referred newborns were more likely to be extremely preterm (<28 weeks; 6.1% vs 3.1%, p<0.001) or very preterm (28–31 weeks; 14.0% vs 8.6%, p<0.001) compared with those inborn. They were also more extremely LBW (<1,000g; 4.6% vs 2.6%, p<0.001) and very LBW (1,000–1,499g; 13.3% vs 6.8%, p<0.001).

**Table 1.** Characteristics of inborn and referred newborns (n=130,773).

|  | Total | Inborn<br>n (%) | Referred<br>n (%) | P-value<br>(Inborn vs<br>Referred) |
| --- | --- | --- | --- | --- |
| **Demographic characteristics** |  |  |  |  |
| **Age at admission** | n=130,773 | n=114,084 | n=16,689 | <0.001 |
| <1 day | 101,480 (77.6) | 91,527 (80.2) | 9,953 (59.6) | <0.001 |
| 1–6 days | 27,103 (20.7) | 21,239 (18.6) | 5,864 (35.1) | <0.001 |
| 7–13 days | 1,466 (1.1) | 932 (0.8) | 534 (3.2) | <0.001 |
| 14–28 days | 724 (0.6) | 386 (0.3) | 338 (2.0) | <0.001 |
| *Median (IQR) days* | *0 (0–0)* | *0 (0–0)* | *0 (0–1)* |  |
| **Sex** | n=130,215 | n=113,609 | n=16,606 | 0.018 |
| Male | 72,804 (55.9) | 63,378 (55.8) | 9,426 (56.7) | 0.018 |
| Female | 57,411 (44.1) | 50,231 (44.2) | 7,180 (43.2) | 0.018 |
| *Missing* | *558* | *475* | *83* |  |
| **Perinatal characteristics** |  |  |  |  |
| **Gestational age** | n=117,736 | n=102,940 | n=14,796 | <0.001 |
| Extremely preterm (<28 weeks) | 4,096 (3.5) | 3,199 (3.1) | 897 (6.1) | <0.001 |
| Very preterm (28–31 weeks) | 10,946 (9.3) | 8,870 (8.6) | 2,076 (14.0) | <0.001 |
| Moderate-to-late preterm (32–36 weeks) | 28,831 (24.5) | 24,999 (24.3) | 3,832 (25.9) | <0.001 |
| Term (37–41 weeks) | 65,608 (55.7) | 58,208 (56.5) | 7,400 (50.0) | <0.001 |
| Late term (≥42 weeks) | 8,255 (7.0) | 7,664 (7.4) | 591 (4.0) | <0.001 |
| *Median (IQR) weeks* | *38 (34–39)* | *38 (35–39)* | *37 (32–39)* |  |
| *Missing* | *13,037* | *11,144* | *1,893* |  |
| **Birthweight, grams (g)** | n=129,785 | n=113,259 | n=16,526 | <0.001 |
| Extremely LBW (<1,000g) | 3,696 (2.8) | 2,937 (2.6) | 759 (4.6) | <0.001 |
| Very LBW (1,000–1,499g) | 9,919 (7.6) | 7,716 (6.8) | 2,203 (13.3) | <0.001 |
| LBW (1,500–2,499g) | 34,254 (26.4) | 29,475 (26.0) | 4,779 (28.9) | <0.001 |
| Normal birthweight (2,500–3,999g) | 73,655 (56.8) | 65,468 (57.8) | 8,187 (49.5) | <0.001 |
| High birthweight (>4,000g) | 8,261 (6.4) | 7,663 (6.8) | 598 (3.6) | <0.001 |
| *Median (IQR) g* | *2800 (2000–3300)* | *2830 (2080–3300)* | *2560(1640–3200)* |  |
| *Missing* | *988* | *825* | *163* |  |
| **Mode of delivery** | n=129,669 | n=113,132 | n=16,537 | <0.001 |
| Normal vertex | 74,784 (57.7) | 61,502 (54.4) | 13,282 (80.3) | <0.001 |
| Vaginal breech | 3,054 (2.4) | 2,632 (2.3) | 422 (2.5) | 0.074 |
| Caesarean delivery | 51,831 (40.0) | 48,998 (43.3) | 2,833 (17.1) | <0.001 |
| *Missing* | *1,104* | *952* | *152* |  |
| **Singleton birth** | n= 117,229 | n=101,414 | n= 15,815 | <0.001 |
| No | 11,376 (9.7) | 9,469 (9.3) | 1,907 (12.1) |  |
| Yes | 105,853 (90.3) | 91,945 (90.7) | 13,908 (87.9) |  |
| *Missing* | *13,544* | *12,670* | *874* |  |
| **Clinical characteristics** |  |  |  |  |
| **Respiratory distress** | n= 122,737 | n=107,138 | n= 15,599 | <0.001 |
| No | 102,614 (83.6) | 91,107 (85.0) | 11,507 (74.8) |  |
| Yes | 20,123 (16.4) | 16,031 (15.0) | 4,092 (26.2) |  |
| *Missing* | *8,036* | *6,946* | *1,090* |  |
| **Oxygen saturation** | n= 98,348 | n=85,335 | n= 13,013 | <0.001 |
| Normal (≥90%) | 82,269 (83.6) | 72,270 (84.7) | 9,999 (76.8) |  |
| Hypoxia (<90%) | 16,079 (16.4) | 13,065 (15.3) | 3,014 (23.2) |  |
| *Missing* | *32,425* | *28,749* | *3,676* |  |
| **Temperature (°C)** | n= 111,837 | n=97,693 | n= 14,144 | <0.001 |
| Normal (36.5–37.5°C) | 56,974 (50.9) | 50,441 (51.6) | 6,533 (46.2) | <0.001 |
| Hypothermia (<36.5°C) | 44,852 (40.1) | 38,996 (39.9) | 5,856 (41.4) | <0.001 |
| Fever (>37.5°C) | 10,011 (9.0) | 8,256 (8.5) | 1,755 (12.4) | <0.001 |
| *Missing* | *18,936* | *16,391* | *2,545* |  |
| **Maternal characteristics** |  |  |  |  |
| **Maternal age** | n=118,775 | n=104,456 | n=14,319 | <0.001 |
| <18 years | 4,421 (3.7) | 3,522 (3.4) | 899 (6.3) | <0.001 |
| 18–24 years | 49,552 (41.7) | 43,394 (41.5) | 6,158 (43.0) | <0.001 |
| 25–34 years | 49,413 (41.6) | 43,852 (42.0) | 5,561 (38.8) | <0.001 |
| 35–39 years | 12,122 (10.2) | 10,771 (10.3) | 1,351 (9.4) | 0.001 |
| ≥40 years | 3,267 (2.7) | 2,917 (2.8) | 350 (2.4) | 0.017 |
| *Median (IQR) years* | *25 (21–31)* | *25 (22–31)* | *25 (21–30)* |  |
| *Missing* | *11,998* | *9,628* | *2,370* |  |
| **Mother alive** | n=90,963 | n=77,906 | n=13,057 | 0.294 |
| No | 586 (0.6) | 493 (0.6) | 93 (0.7) |  |
| Yes | 90,377 (99.4) | 77,413 (99.4) | 12,964 (99.3) |  |
| *Missing* | *39,810* | *36,178* | *3,632* |  |
| **Mother HIV status** | n=122,571 | n=108,014 | n=14,557 | 0.001 |
| Negative | 117,233 (95.6) | 103,389 (95.7) | 13,844 (95.1) |  |
| Positive | 5,338 (4.4) | 4,625 (4.3) | 713 (4.9) |  |
| *Missing* | *8,202* | *6,070* | *2,132* |  |

**Abbreviations:** SD, standard deviation; IQR, interquartile range; LBW, low birth weight; °C, degree Celsius; HIV, human immunodeficiency virus

Clinically, referred newborns presented more frequently with respiratory distress (26.2% vs 15.0%, p<0.001), hypoxia (oxygen saturation <90%; 23.2% vs 15.3%, p<0.001) and fever (temperature >37.5^0^C; 12.4% vs 8.5%, p<0.001). Maternal characteristics varied slightly.

Among referred newborns, 6.3% were born to mothers younger than 18 years, compared with 3.4% among inborn newborns (p<0.001). Among mothers with known HIV status, HIV prevalence was slightly higher among those of referred newborns (4.9% vs 4.3%, p=0.001).

#### Objective 2: Clinical diagnoses

Clinical diagnoses were reported for 126,038 (96.4%; 126,038/130,773), including 109,915 inborn and 16,123 referred newborns (**Figure 1**). Prematurity was the most common diagnosis overall, accounting for 38.4% of admissions and was more frequent among referred (48.8%) than inborn (36.8%) newborns. Intrapartum-related complications were the second most common diagnosis, affecting 22.0% of inborn and 20.1% of those referred.

**Figure 1.**
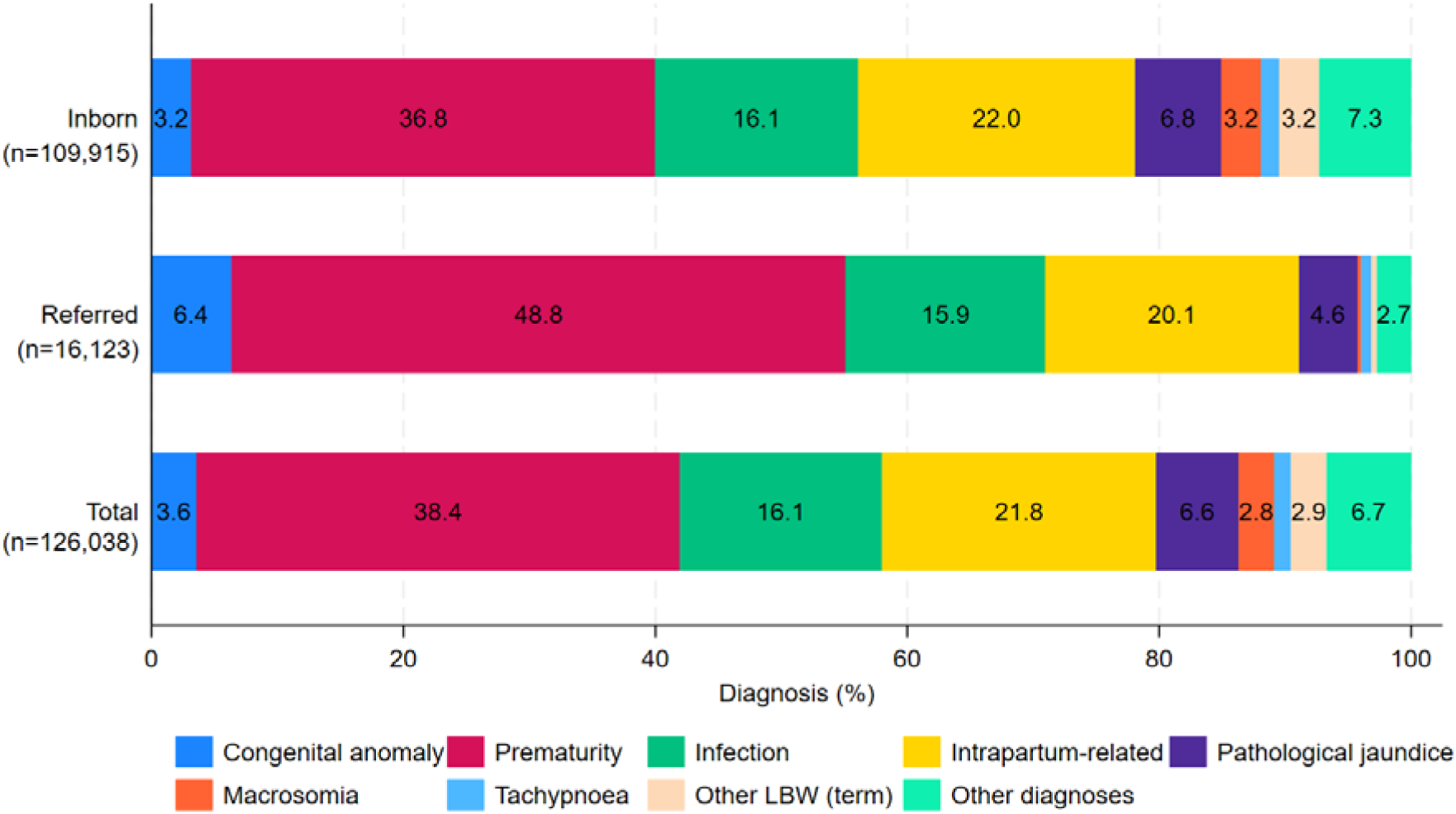
**Clinical diagnoses among inborn and referred newborns (n=126,038).** Congenital anomalies accounted for 3.6% of all diagnoses and were twice as common among referred (6.4%) than inborn (3.2%) newborns. Pathological jaundice occurred in 6.6% of admissions overall and was slightly higher among inborn (6.8%) than referred (4.6%) newborns. Macrosomia (2.8%) and tachypnoea (1.3%) were relatively uncommon. Other categories of LBW at term represented 2.9% of all admissions. The remaining diagnoses included conditions such as newborns admitted for observation, feeding difficulties, dehydration, rhesus isoimmunization, and other less frequent causes.

Infections were reported in 16.1% of newborns, with similar proportions among inborn (16.1%) and those referred (15.9%).

#### Objective 3: Factors associated with mortality

A total of 17,735 newborns died among 130,676 with mortality outcome data, representing an overall unadjusted mortality rate of 13.6% (95% CI 13.4–13.8). Referred newborns had higher mortality rate (29.0%, 95% CI: 28.3–29.7) compared to inborn newborns (11.3%, 95% CI: 11.1–11.5) (p<0.001). This was also observed in univariable analysis, with 3 times higher odds of mortality among referred compared to inborn newborns (OR 3.20, 95% CI 3.08–3.33) **(Table 2).**

**Table 2:** Association between birth location and newborn mortality (n=130,676).

|  | n | Died (row %) | OR (95% CI) | P-value |
| --- | --- | --- | --- | --- |
| Inborn | 114,003 | 12,900 (11.3) | 1 | <0.001 |
| Referred | 16,673 | 4,835 (29.0) | 3.20 (3.08– 3.33) |  |

n=number; OR=odds ratio; CI=confidence interval

Table 3 presents the univariable analysis of factors associated with mortality among all newborns, those inborn, and referred. Mortality did not differ by sex overall, however among referred newborns, females had slightly higher odds of death (OR 1.07, 95% CI 1.00–1.15). In all newborns, being extremely LBW had the highest odds of death (OR 56.00, 95% CI 51.16–61.31) followed by extremely preterm (OR 24.10, 95% CI 22.44–25.88). Preterm non-SGA (OR 3.21, 95% CI 3.08–3.34), and preterm SGA (OR 4.21, 95% CI 3.97–4.47) had higher odds of death compared to term non-SGA newborns. This was also observed among inborn and referred groups, although the strength of association varied.

**Table 3.**
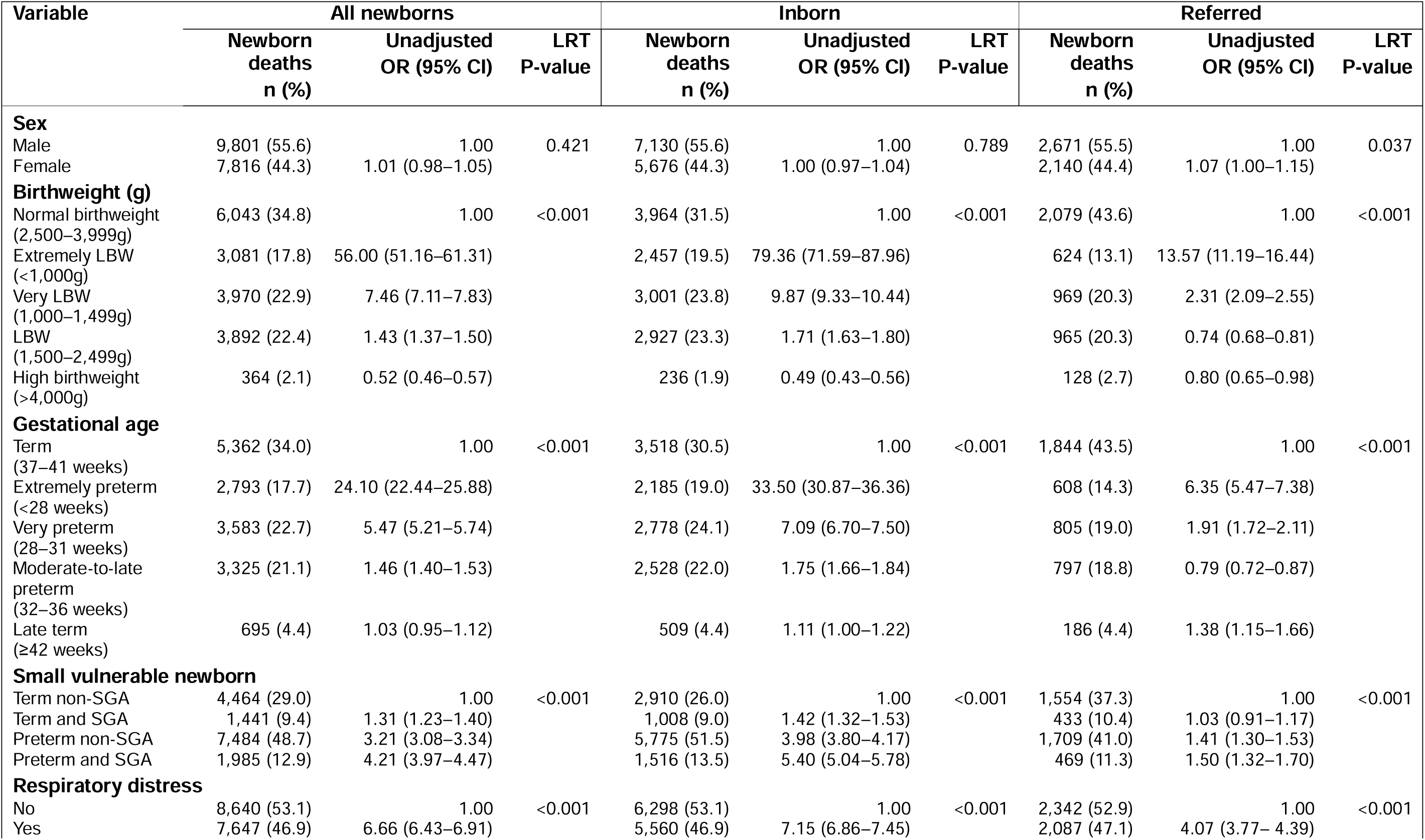
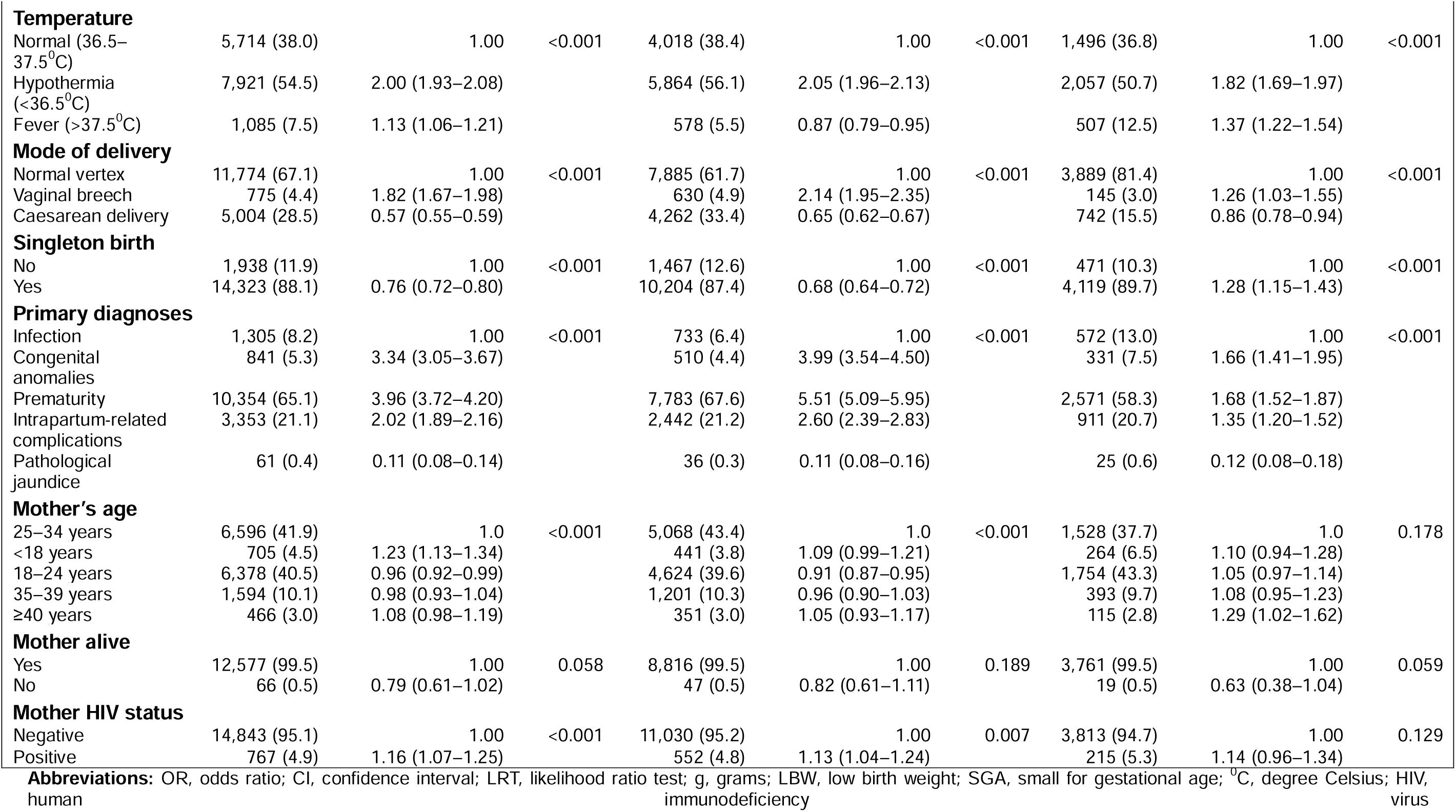
Unadjusted univariable analysis of factors associated with mortality among inborn and referred newborns.

There was strong evidence that having respiratory distress was associated with 7 times higher odds of death among inborn (OR 7.15, 95% CI 6.86–7.45) and 4 times higher odds among referred newborns (OR 4.07, 95% CI 3.77–4.39). Hypothermia was associated with almost twice the odds of mortality compared to normal temperature among inborn (OR 2.05, 95% CI 1.96–2.13) and referred newborns (OR 1.82, 95% CI 1.69–1.97). Compared to newborns diagnosed with infection, those with congenital anomalies, prematurity or intrapartum-related complications were strongly associated with increased odds of death in both groups. In contrast, pathological jaundice had reduced odds of death. The multivariable logistic regression of factors independently associated with neonatal mortality are summarised in **Additional File 2**.

## DISCUSSION

Neonatal mortality differed markedly between inborn and referred newborns. Analysis of newborn records also showed disparities in their characteristics and morbidity outcomes, which may suggest persistent gaps in perinatal care, referral processes, and timely recognition of complications.

The overall unadjusted mortality rate of 11.3% among those inborn was comparable to 10.2% reported in a group of inborn newborns in Kenya [11]. Referred newborns had a higher unadjusted mortality rate of 29.0% which was similar to findings in Ethiopia (30.6%) and Uganda (33.0%) [22, 33]. The higher mortality rate observed among those referred is likely due to a combination of baseline risk factors that disproportionately affect this group [10], as well as delays in accessing essential care throughout the referral process [33]. For example, while time of referral was not captured, majority of hospitals implementing with NEST360 in Kenya are in semi-urban and urban areas and travel time may be increased due to traffic congestion. This has been found as a barrier to accessing emergency maternal and neonatal care in other countries in sub-Saharan Africa [21, 34].

Among referred newborns, significant predictors of mortality included birthweight, gestational age, respiratory distress, oxygen saturation, temperature, and diagnoses. Newborns with extremely LBW are often more vulnerable to complications, especially respiratory distress syndrome and infections, reducing their probability of survival [35, 36]. The associations between respiratory distress, hypothermia, and fever with mortality likely indicates the burden of prematurity in this group. Compared with referred newborns diagnosed with infection, those with congenital anomalies had 66% higher odds of death, while those with intrapartum-related complications had 35% higher odds. Congenital anomalies are a well-documented cause of newborn mortality [37]. The association of intrapartum complications with mortality might be related to their acute, often unexpected onset, limited time for diagnosis and delays in referral [38]. Unanticipated complications, such as adverse birth events like severe preterm birth, neonatal encephalopathy following intrapartum events, or major congenital anomalies (in the absence of antenatal ultrasound), offer less lead time for preparation before a referral becomes crucial [39, 40]. In contrast, pathological jaundice was associated with reduced odds of death, likely indicating that these newborns are detected early and responsive to phototherapy.

Timing of admission also differed between the groups, with inborn newborns more likely to be admitted within 24 hours of birth (80.2%) than those referred in (59.6%). Delayed recognition of neonatal illness due to inadequate infrastructure at lower-level facilities, or challenges in timely transfer may contribute to poorer outcomes particularly for newborns requiring specialised care [41, 42]. This emphasises the need for rapid referral and improved quality of care during the first day of life, which is a ‘critical period’ for newborn survival [43].

### Strengths and limitations

The study has several strengths. Analyses leveraged the NID, which is a large clinical database that included data on newborns admitted to 13 hospitals across 10 of the 47 counties in Kenya [27]. This represents diverse healthcare settings and is useful for external validity and generalisability of findings. The large sample size enabled robust estimation of mortality risks and sufficient statistical power to examine relatively uncommon but important outcomes such as congenital anomalies. Use of routinely collected data also strengthens relevance to practice as they reflect care delivered under routine clinical conditions rather than ideal conditions in studies. In addition, unique comparison of inborn and referred newborns provides insight into health system performance across levels of care including access to specialised services rather than solely on individual hospital performance.

However, there are various limitations that should be considered. First, only the main diagnosis was analysed for each newborn. SSNBs frequently present with multiple concurrent conditions and this approach could have resulted in underestimation of contribution of comorbidities to mortality. Second, residual confounding is possible as important factors such as maternal health, antenatal care quality, socio-economic status, or transport quality were not fully captured and may partially explain observed differences.

Selection bias is also likely, as referred newborns likely represent a higher-risk population and comparisons with those inborn could be biased [25]. Third, the data did not include level of referring facilities hence assessment of variation in referral patterns by level of care was not conducted. In addition, clinical practices and resource availability were not standardised across hospitals and variation in staffing, hospital capacity or adherence to protocols could have affected outcomes, independent of referral status [44]. Fourth, disruptions in service delivery such as health worker strikes may have influenced referral and newborn outcomes. Although such events were not systematically captured and are not expected to have been frequent, they represent a potential source of unmeasured variation in care. Finally, temporal trends in mortality and referral patterns were not explicitly examined and could have changed over time especially with ongoing NEST360 implementation and other health system interventions.

#### Implications for policy and practice

The gap in mortality between inborn and referred newborns shows the need to strengthen neonatal referral systems. Policy efforts should prioritise clear criteria for SSNB referral, improved coordination between facilities, and early maternal assessment to refer complicated pregnancies in a timely manner when needed [45]. Assessing facility readiness and processes for managing SSNBs would support efforts to improve outcomes through more effective referral and admission systems [46]. A hub-and-spoke model may also be a useful approach to connect lower levels of care (spokes) with tertiary facilities (hubs) for capacity building and oversight. This model has shown promise in supporting quality improvement across facilities in India [47], and in improving access to cervical cancer screening in rural Ghana [48].

Improving quality of neonatal transport and early interventions is also vital to reduce preventable newborn mortality of referred babies. This could include pre-transfer stabilisation at peripheral facilities, standardised referral protocols, training of transport teams, and prompt triaging on arrival at the receiving facilities [6, 10, 19]. Many of these processes can be monitored using routinely collected hospital data which provide an opportunity for ongoing quality improvement. Experiences from NEST360 have exemplified the value of clinical data in supporting decision-making and guiding quality improvements, which were associated with reductions in hospital neonatal mortality [49].

#### Implications for future research

Future research is needed to better understand pathways contributing to excess mortality among referred newborns. Studies incorporating referral-to-admission times, modes of transport, and quality of pre-transfer stabilisation could help identify critical delays and modifiable gaps in care. Analysis models that allow for multiple concurrent diagnoses could also better examine the complexity of neonatal conditions. Linking neonatal admissions data with maternal records may improve understanding of antenatal risk factors and opportunities for earlier referral. In addition, follow-up studies post-discharge are needed to evaluate long term outcomes beyond the inpatient period.

## Conclusions

There was substantial variation in characteristics and outcomes between inborn and referred newborns. Referred babies were more likely to be premature, LBW, and clinically unstable, which may in part indicate selection bias, as those at greater risk are more likely to be referred. However, the higher mortality seen could suggest that this inherent vulnerability may be compounded by gaps in continuum of care including delayed referral, inadequate pre-referral stabilisation, and limited capacity for SSNC at lower-level hospitals. The high prevalence of intrapartum-related complications among this group also shows need for earlier identification of complicated pregnancies and timely maternal transfer before delivery. Implementation of evidence-based practices is essential to increase coverage to specialised care and to help newborns survive and thrive.

## LIST OF ABBREVIATIONS

aOR: Adjusted odds ratio
CI: Confidence interval
CIN: Clinical Information Network
EmONC: Emergency Obstetric and Newborn Care
EWENE: Every Woman, Every Newborn, Everywhere
HIV: Human immunodeficiency virus
IQR: Interquartile range
KEMRI: Kenya Medical Research Institute
LBW: Low birthweight
LMICs: Low- and middle-income countries
LRT: Likelihood ratio test
LSHTM: London School of Hygiene & Tropical Medicine
NEST360: Newborn Essential Solutions and Technologies 360
NID: Neonatal Inpatient Dataset
NMR: Neonatal mortality rate
REDCap: Research Electronic Data Capture
SDG: Sustainable Development Goal
SERU: Scientific and Ethics Review Unit
SGA: Small for gestational age
SSNB: Small and sick newborn
SSNC: Small and sick newborn care
WHO: World Health Organisation

## DECLARATIONS

### Ethics and consent to participate

Ethical approvals for NEST360 were granted by London School of Hygiene & Tropical Medicine (LSHTM) Research Ethics Committees (Ethics Ref: 21892), Kenya Medical Research Institute (KEMRI) Scientific and Ethics Review Unit (SERU) (SERU number: 3459), and Rice University. All patient-level data were de-identified.

### Consent for publication

Not applicable.

### Availability of data and material

Data sharing and transfer agreements were jointly developed and signed by all collaborating partners in the NEST360 alliance. The dataset generated during the current study will be available upon request subject to approval by the NEST360 learning network and collaborating parties. The NID tool, data dictionary, and associated materials are available from the Newborn Toolkit for Small and Sick Newborn Care (https://www.newborntoolkit.org) and NEST360 website (https://nest360.org).

## Funding

This work is funded through the NEST360 Alliance with thanks to John D. and Catherine T. MacArthur Foundation, the Bill & Melinda Gates Foundation, ELMA Philanthropies, The Children’s Investment Fund Foundation UK, The Lemelson Foundation, the Sall Family Foundation, and the Ting Tsung and Wei Fong Chao Foundation under agreements to William Marsh Rice University.

### Authors’ contributions

This work was done in partnership with the NEST360 Alliance. All authors were involved in initial conceptualization and design of the study. JB, SMC, and REP with WM developed detailed research questions and overall analysis plan for this paper. These were refined with input from the wider team. JB conducted formal analysis and drafted the initial manuscript. REP assisted with data management and statistical analysis. The manuscript underwent review and revision by all authors. All authors reviewed and gave their consent to the final version of the manuscript. The authors’ views are their own, and not necessarily from any of the institutions they represent.

## Data Availability

Data sharing and transfer agreements were jointly developed and signed by all collaborating partners in the NEST360 alliance. The dataset generated during the current study will be available upon request subject to approval by the NEST360 learning network and collaborating parties. The Neonatal Inpatient Dataset (NID) tool, data dictionary, and associated materials are available from the Newborn Toolkit for Small and Sick Newborn Care (https://www.newborntoolkit.org) and NEST360 website (https://nest360.org).

## Acknowledgements

Firstly, and most importantly, we thank the women, families, and newborns together with all the health workers involved in this work. We also thank KEMRI-Wellcome Trust Research Programme, the Clinical Information Network, and NEST360, and all the data teams involved in collecting the data. We also would like to thank the Ministry of Health in Kenya.

## ADDITIONAL FILES

**Additional file 1:** Flow-chart of inborn and referred newborns sample inclusion (n=130,773).

**Additional file 2.** Multivariable logistic regression of factors associated with mortality (n=72,900).

