## Additional files for "Clinical outcomes and mortality risk among inborn and referred newborns admitted to hospitals in Kenya"

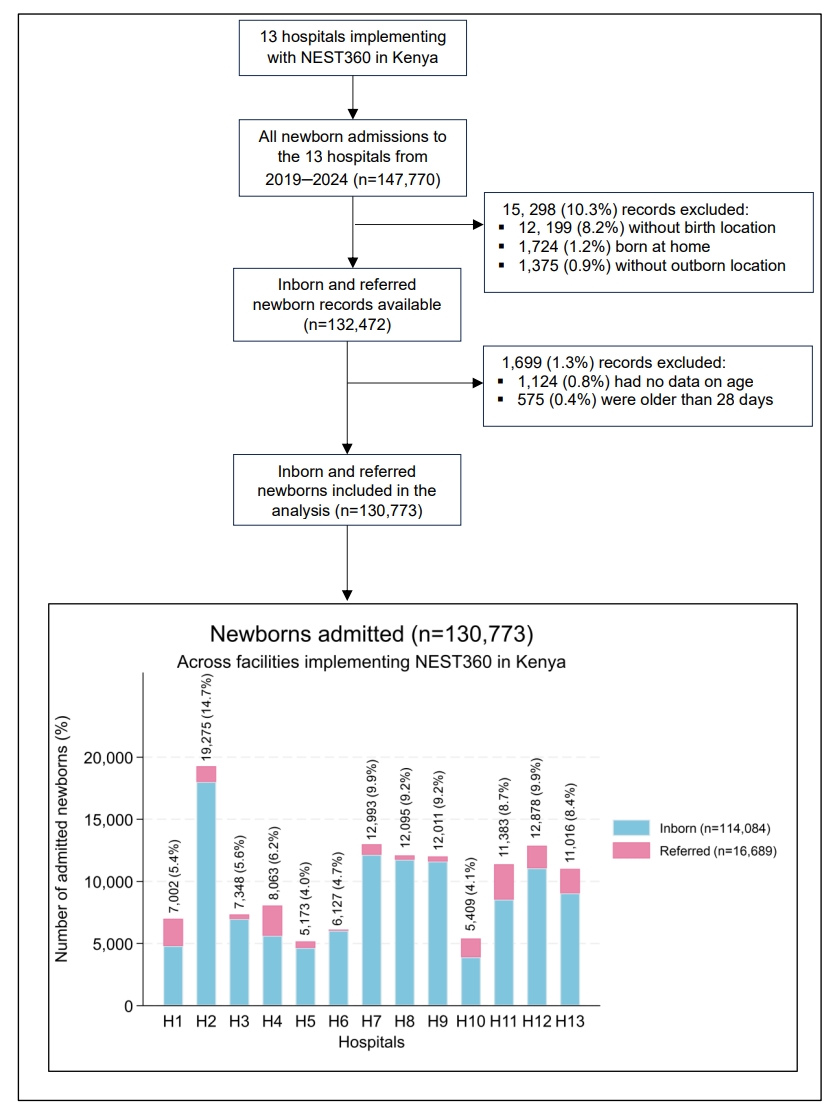

**Additional file 1. Flow-chart of inborn and referred newborns sample inclusion (n=130,773).**

**Additional file 2. Multivariable logistic regression of factors associated with mortality (n=72,900).**

| **Variable** | **Adjusted OR (95% CI)** | **LRT P-value** |
| --- | --- | --- |
| **Birth location** |  |  |
| Inborn | 1.00 | <0.001 |
| Referred | 2.54 (2.39–2.71) |  |
| **Sex** |  |  |
| Male | 1.00 | 0.013 |
| Female | 0.94 (0.89–0.99) |  |
| **Birthweight (g)** |  |  |
| Normal birthweight (2,500–3,999g) | 1.00 | <0.001 |
| Extremely LBW (<1,000g) | 18.45 (15.71–21.66) |  |
| Very LBW (1,000–1,499g) | 3.41 (3.06–3.80) |  |
| LBW (1,500–2,499g) | 1.08 (1.00–1.17) |  |
| High birthweight (>4,000g) | 0.89 (0.76–1.04) |  |
| **Gestational age** |  |  |
| Term (37–41 weeks) | 1.00 | <0.001 |
| Extremely preterm (<28 weeks) | 2.56 (2.22–2.95) |  |
| Very preterm (28–31 weeks) | 1.20 (1.08–1.34) |  |
| Moderate-to-late preterm (32–36 weeks) | 0.79 (0.73–0.86) |  |
| Late term (≥42 weeks) | 1.17 (1.05–1.30) |  |
| **Respiratory distress** |  |  |
| No | 1.00 | <0.001 |
| Yes | 4.43 (4.20–4.67) |  |
| **Temperature** |  |  |
| Normal (36.5–37.5^0^C) | 1.00 | <0.001 |
| Hypothermia (<36.5^0^C) | 1.28 (1.22–1.35) |  |
| Fever (>37.5^0^C) | 1.44 (1.30–1.59) |  |
| **Mode of delivery** |  |  |
| Normal vertex | 1.00 | <0.001 |
| Vaginal breech | 1.19 (1.03–1.36) |  |
| Caesarean delivery | 0.89 (0.84–0.94) |  |
| **Diagnosis** |  |  |
| Infection | 1.00 | <0.001 |
| Congenital anomaly | 2.75 (2.41–3.13) |  |
| Prematurity | 1.65 (1.50–1.81) |  |
| Intrapartum-related complication | 1.96 (1.78–2.14) |  |
| Pathological jaundice | 0.16 (0.11–0.22) |  |
| **Mother’s age** |  |  |
| 25–34 years | 1.00 | 0.003 |
| <18 years | 1.11 (0.98–1.26) |  |
| ≥40 years | 1.32 (1.14–1.53) |  |
| **Mother HIV status** |  |  |
| Negative | 1.00 | 0.026 |
| Positive | 1.15 (1.02–1.29) |  |

**Abbreviations:** OR, odds ratio; CI, confidence interval; LRT, likelihood ratio test; g, grams; LBW, low birth weight; ^0^C, degree Celsius; HIV, human immunodeficiency virus
